# Analysis of whole genome sequence data shows association of Alzheimer’s disease with rare coding variants in *ABCA7, PSEN1, SORL1* and *TREM2*

**DOI:** 10.1101/2025.07.27.25332260

**Authors:** David Curtis, Shujaani Joseph, the Alzheimer’s Disease Neuroimaging Initiative

## Abstract

Previous studies have reported associations between risk of Alzheimer’s disease (AD) or dementia and rare coding variants in a number of genes. A two stage strategy was used in which a previously released whole exome sequenced sample was used to prioritise 100 genes showing the strongest evidence for association with AD. These genes were then analysed in a newly released whole genome sequenced sample to identify those which showed statistically significant evidence for rare coding variant association. Association analysis of loss of function (LOF) and nonsynonymous variants was carried out in 18,998 protein coding genes using 11,188 controls and 5,808 cases, with nonsynonymous variants being annotated using 45 different pathogenicity predictors. The 100 genes showing strongest evidence for association were then analysed in a new sample of 27,749 controls and 13,234 cases using only the pathogenicity predictor which had performed best in the first sample. Four genes were statistically significant after correction for multiple testing: *ABCA7, PSEN1, SORL1* and *TREM2*. The association of different categories of variant with AD was characterised and the pattern was seen to vary between genes. This study quantifies the contribution of different types of variant within each gene to AD risk. In general these variants are probably too rare to be clinically useful for assessing individual risk of AD. Further research into the mechanisms whereby the products of these genes affect AD pathogenesis may aid development of novel therapeutic strategies.

## Introduction

A number of genetic risk factors can modify risk of dementia in general and Alzheimer’s disease (AD) in particular and understanding their contribution may provide insights into mechanisms of pathogenesis and assist the development of therapeutic interventions [1–3]. Rare coding variants in *APP, PSEN1* and *PSEN2* are causally involved in early onset AD while rare coding variants in *PSEN1* have also been shown to be associated with the common, late onset form of AD [2,4]. A rare nonsynonymous variant in *TREM2*, rs75932628-T producing an arginine to histidine substitution (R47H), was reported to be associated with late onset AD with OR (95% CI) 2.92 (2.09 - 4.09) [5]. Other studies have identified rare variants in *SORL1, ABCA7, ATP8B4*, and *ABCA1* as being also implicated in AD susceptibility [6]. More recently, a gene-based exome-wide association study (ExWAS) of White British UK Biobank participants with a phenotype described as clinically diagnosed/proxy AD and related dementia (ADRD) found associations for ten genes significant after FDR correction: *SORL1, GRN, PSEN1, ABCA7, GBA, ADAM10, FRMD8, DDX1, DNMT3L* and *MORC1* [7]. Another study which used all exome-sequenced UK Biobank participants with and a phenotype of personal or family history of dementia found association with rare coding variants in *TREM2, SORL1* and *ABCA7* [8].

The Alzheimer’s Disease Sequencing Project (ADSP) has released whole genome sequence (WGS) data for 43,795 cases and controls as part of NIAGADS, which is the National Institute on Aging (NIA) designated national data repository for AD and AD-related dementia (ADRD) human genetics research [9]. Here we report the results for analyses of rare coding variants in 100 genes nominated by results obtained in the previously released exome-sequenced sample of 16,996 cases and controls.

## Methods

Data used in the preparation of this article was obtained from the Alzheimer’s Disease Neuroimaging Initiative (ADNI). The ADNI was launched in 2003 as a public-private partnership, led by Principal Investigator Michael W. Weiner, MD. The primary goal of ADNI has been to test whether serial magnetic resonance imaging, positron emission tomography, other biological markers, and clinical and neuropsychological assessment can be combined to measure the progression of mild cognitive impairment and early Alzheimer’s disease. The investigators within the ADNI contributed to the design and implementation of ADNI and/or provided data but did not participate in analysis or writing of this report [10]. A complete listing of ADNI investigators can be found at: http://adni.loni.usc.edu/wp-content/uploads/how_to_apply/ADNI_Acknowledgement_List.pdf.

Phenotype information and variant calls using the GRCh38 assembly for case-control subjects of the Alzheimer’s Disease Sequencing Project (ADSP) were downloaded from the NIAGADS website (https://www.niagads.org/adsp/content/study-design). Ethical approval and informed consent had been obtained by the researchers who generated this dataset and ethics approval for the analyses performed was obtained from the UCL Research Ethics Committee (11527/003). As described previously, participants were at least 60 years old and although different diagnostic procedures were used by different contributing studies all cases met NINCDS-ADRDA criteria for possible, probable or definite AD based on clinical assessment, or had presence of AD (moderate or high likelihood) upon neuropathology examination [11]. The phenotype files were downloaded and a list of genetically unique samples with harmonized phenotypes for each sample was generated using the recommended script provided by NIAGADS (ADSPIntegratedPhenotypes_Script_2023.08.08.R). These files were also used to obtain the number of *APOE ε*3 and ε4 alleles carried by each subject.

The data was analysed in two phases. The first phase, to screen all genes for evidence of association, utilised previously released whole exome sequence data for 16,996 cases and controls with the genetic data being obtained using standard methods as described previously (ng00067.v2) [12] [13]. As described below, this dataset was used to nominate the 100 genes showing the strongest evidence for association of rare coding variants with AD. For the second phase, these 100 genes were analysed in the newly released Alzheimer’s Disease Sequencing Project Release 4 sample of 43,795 cases and controls which had undergone whole genome sequencing [14] [15].

The methods used were essentially the same as those applied to the UK Biobank dataset, with the description repeated here for convenience [8]. To test for association with rare coding variants, attention was confined to loss of function (LOF) and nonsynonymous variants with minor allele frequency (MAF) < 0.01. All exonic variants were annotated using Variant Effect Predictor (VEP) [16]. Nonsynonymous variants were additionally annotated using the AlphaMissense plug-in of VEP, which produces a raw score and a categorisation of likely pathogenic, likely benign or ambiguous, these three categories being converted to numerical scores of 2, 0 or 1 respectively [17]. Additionally, rank prediction scores were obtained for an additional 43 different predictors of pathogenicity as provided for all possible nonsynonymous variants in dbNSFP v4 [18].

In order to carry out weighted burden analysis, for each LOF or nonsynonymous variant a score was defined. For LOF variants this score used a parabolic function of allele frequency as previously described, with extremely rare variants (MAF ∼ 0) being assigned a score of 10 and less rare variants (MAF=0.01) being assigned a score of 1 [19,20]. For nonsynonymous variants, this frequency score was assigned in the same way but was then multiplied by the score for a pathogenicity predictor in order to produce an overall score for that variant. For each gene, each participant would receive a LOF score and a nonsynonymous score, consisting of the sums of the scores for these classes of variant carried by that participant. To test for association, multiple linear regression was performed using sex, 20 principal components and *APOE ε*3 and ε4 doses as covariates. For the null model, AD status was predicted solely from these covariates. For the alternative model, AD status was predicted from these covariates along with the LOF and nonsynonymous scores. The likelihoods of these models were compared and a p value obtained assuming that twice the log likelihood difference followed a chi-squared distribution with two degrees of freedom and the statistical evidence for association was summarised as minus log10(p) (MLP).

Because previous work had shown that LOF variants and nonsynonymous variants could make different relative contributions to disease risk and because different predictors of pathogenicity had differential performance across different genes, a two-stage approach to analysis was followed [21]. In the first stage, the dataset of WES participants was used to carry out 45 different regression analyses as described above for each protein-coding gene using each of the 45 pathogenicity predictors. This yielded 45 MLPs for each gene, the maximum being denoted MaxMLP. In the second stage, the WGS participants were used for analyses of only the 100 genes showing the strongest evidence for association in the first phase and each gene was analysed using only the predictor which had produced the maximum MLP for that gene in the first phase. Thus only 100 tests were performed in the second phase, meaning that a gene could be declared statistically significant if it produced a test statistic significant at p < 0.05/100. In the second stage analysis, any WGS participants who had been included in the WES sample were excluded. Additionally, if for any of the 100 genes there was no signal for association with LOF variants (Wald statistic for LOF score > 0.05) then the second stage analysis was carried out using only the nonsynonymous variant score, resulting in a test with only 1 degree of freedom.

In the second stage analysis, *PSEN1* produced very strong evidence for association with MLP = 34.88. Closer examination of these results showed that the signal was driven in part by the nonsynonymous variant chr14:73192712 G>C, for which the C allele was observed in 62 cases and only 5 controls. This is rs63750082, a well-known cause of familial Alzheimer’s disease in people with Caribbean-Hispanic ancestry [22]. In order to explore the effects of other variants in this gene, the analyses were repeated with the rs63750082 genotypes excluded.

Similarly, *TREM2* initially produced evidence for association with MLP = 8.86. Examination of the results revealed that this was partially driven by the previously reported variant chr6:41161514C>T, which is rs75932628, and which had MAF 0.0027 in controls and 0.0060 in cases. It was noted that another nearby variant, 6:41161469C>T or rs143332484, was not extremely rare and had MAF 0.0077 in controls and 0.0110 in cases. In order to better assess the effect of other variants in *TREM2*, analyses were performed with these variants excluded. In addition, in order to explore their contribution to AD risk they were both entered into a logistic regression analysis along with sex, principal components and *APOE* allele doses as covariates.

For all genes producing statistically significant evidence for rare variant association in the second phase, subsidiary analyses were carried out in using all variants with MAF < 0.01 using VEP to group them broadly into functional categories: LOF, protein-altering, synonymous, indel etc, 5’ UTR, 3’ UTR, intronic etc and splice region. Logistic regression analyses were carried out using the raw counts of variants in each category, unweighted by MAF, and with variants additionally categorised according to whether or not they had a predictor score greater than a threshold value of 0.35.

As a secondary analysis, for any of the genes listed above as previously reported to be associated with AD and/or dementia but not attaining statistical significance in this study, association testing was carried out in the WGS sample using the LOF score and the score for the best performing predictor. Additionally, the raw counts for different variant categories were analysed in the same way as for the statistically significant genes.

## Results

The WES sample consisted of 11,188 controls and 5,808 cases. Results were obtained for 18,998 protein coding genes. The 100 maximum MLPs ranged from 15.6 for *TREM2* to 3.66 for *ICA1L*. Full results for all genes and all predictors are contained in Supplementary Table 1. The 100 genes with the highest maximum MLPs were taken forward for analysis in the WGS sample.

After excluding subjects present in the WES sample, the WGS sample consisted of 27,749 controls and 13,234 cases. Of the 100 genes tested in the WGS, four produced results which were statistically significant after correction for multiple testing: *ABCA7, PSEN1, SORL1* and *TREM2*. The results for these genes in the first and second stage analyses, along with the pathogenicity predictor used, are shown in Table 1. Results for all 100 genes are shown in Supplementary Table 2.

**Table 1.** For each gene achieving statistical significance in the second stage analysis, the table shows the maximum −log10(p) (MLP) produced by any of 45 pathogenicity predictors in the first stage analysis of 16,996 exome-sequenced participants along with the predictor producing this MLP and the results obtained in the second stage using 40,983 participants. All results are obtained from multiple logistic regression analyses of AD with sex, 20 principal components and *APOE* genotypes as covariates. For *ABCA7* and *SORL t*he MAF-weighted LOF scores and pathogenicity scores were both included but for *PSEN1* and *TREM2* the LOF scores were not significantly associated in the first stage analysis so only the MAF-weighted pathogenicity scores were included in the second stage. For *PSEN1*, rs63750082 genotypes were excluded from the WGS analysis. For *TREM2*, rs75932628 and rs143332484 genotypes were excluded from the WGS analysis.

| Gene | MaxMLP (WES) | Predictor | MLP (WGS) |
| --- | --- | --- | --- |
| *ABCA7* | 10.37 | fathmm_XF_coding | 7.51 |
| *PSEN1* | 4.82 | phyloP17way_primate | 15.74 |
| *SORL1* | 7.97 | VEST4 | 3.67 |
| *TREM2* | 15.64 | MetaLR | 4.26 |

The two less rare variants which had been excluded from the *TREM2* analysis, rs75932628 and rs143332484, were together entered into a logistic regression analysis with sex, principal components and *APOE* allele doses as covariates. Both showed independent evidence for association with AD. The previously reported variant, rs75932628, had OR = 1.71 (1.35 - 2.18), p = 8.4 × 10^−6^ and rs143332484 had OR = 1.28 (1.09 - 1.50), p = 0.0023.

In order to characterise the effects on risk of different categories of variant in these genes, logistic regression analysis was carried out on the raw counts of variants in each category as obtained from VEP and Table 2 shows results for counts of LOF variants, protein-altering variants and variants with high pathogenicity scores, along with results for any other category which was nominally significant at p<0.05 for that gene. The full results for all categories are shown in Supplementary Table 3.

**Table 2.** Table showing results of multiple logistic regression analysis of AD in 40,983 participants for statistically significant genes using raw counts of LOF and protein altering variants as well as those variants having a score of > 0.35 for the relevant predictor. Results are also shown for any variant category for which the absolute value of the SLP exceeds 1.3 (equivalent to p < 0.05). For *PSEN1*, rs63750082 genotypes were excluded from the analysis. For *TREM2*, rs75932628 and rs143332484 genotypes were excluded from the analysis.

| Gene / category / predictor | Number of separate variants | Total count in controls | Mean count in controls | Total count in cases | Mean count in cases | OR (95% confidence interval) | SLP |
| --- | --- | --- | --- | --- | --- | --- | --- |
| *ABCA7* |  |  |  |  |  |  |  |
| LOF | 163 | 1444 | 0.05212 | 808 | 0.06119 | 1.23 (1.12-1.35) | 5.37 |
| Protein altering | 813 | 4105 | 0.14814 | 1990 | 0.15066 | 0.98 (0.88-1.08) | -0.19 |
| fathmm_XF_coding | 429 | 2124 | 0.07666 | 1119 | 0.08473 | 1.34 (1.08-1.67) | 2.21 |
| Intronic etc | 69 | 1259 | 0.04555 | 305 | 0.02315 | 0.83 (0.72-0.96) | -2.09 |
| *PSEN1* |  |  |  |  |  |  |  |
| LOF | 10 | 7 | 0.00025 | 9 | 0.00068 | 1.80 (0.65-4.96) | 0.61 |
| Protein altering | 126 | 289 | 0.01042 | 225 | 0.01702 | 1.02 (0.81-1.30) | 0.08 |
| phyloP17way_primate | 71 | 68 | 0.00245 | 101 | 0.00763 | 5.63 (3.26-9.73) | 9.58 |
| *SORL1* |  |  |  |  |  |  |  |
| LOF | 43 | 125 | 0.00451 | 98 | 0.00741 | 1.44 (1.08-1.93) | 1.90 |
| Protein altering | 616 | 113360 | 4.08659 | 54219 | 4.09860 | 1.04 (0.95-1.13) | 0.42 |
| VEST4 | 475 | 1912 | 0.06892 | 1024 | 0.07740 | 1.10 (0.92-1.31) | 0.54 |
| Splice region | 65 | 171 | 0.00616 | 116 | 0.00877 | 1.33 (1.03-1.72) | 1.60 |
| *TREM2* |  |  |  |  |  |  |  |
| LOF | 16 | 23 | 0.00083 | 20 | 0.00151 | 1.58 (0.83-3.00) | 0.81 |
| Protein altering | 69 | 595 | 0.02144 | 303 | 0.02290 | 1.11 (0.79-1.54) | 0.26 |
| MetaLR | 29 | 393 | 0.01416 | 205 | 0.01550 | 2.40 (0.98-5.84) | 1.30 |
| Three prime UTR | 42 | 764 | 0.02754 | 314 | 0.02373 | 1.25 (1.08-1.45) | 2.59 |

Table 2 shows that each of the four genes has a somewhat different pattern of effects which contribute to the overall evidence for association. One thing to note is that the predictor which had produced the best evidence for association in the WES sample is different for each gene. There is no gene for which the overall category of “Protein altering” (mostly nonsynonymous variants) is associated with higher risk but for *ABCA7, PSEN1* and *TREM2* there is evidence for association with the subset of variants which score highly with the relevant pathogenicity predictor. Another feature is that these results generally have weaker statistical significance than those obtained for the main analyses, presumably reflecting the lack of weighting for allele frequency and the conversion of raw predictor scores to dichotomised scores.

For *ABCA7* the main signal comes from LOF variants, which have an overall allele count of over 2,000 and which produce an SLP of 5.37 though are associated with only a modest increase in risk, with OR = 1.23 (1.12-1.35). There is possibly some additional signal from variants which score highly with the fathmm_XF_coding predictor, with SLP = 2.21 and OR = 1.34 (1.08-1.67). However intronic variants as a category are associated with somewhat reduced risk, with SLP = −2.09 and OR = 0.83 (0.72-0.96).

The situation with *PSEN1* is very different. Here, LOF variants are extremely rare and do not yield any evidence for association. While in general protein altering variants have no overall effect on risk, those with a high score with the phyloP17way_primate produce very strong evidence for association (SLP = 9.58) with a large average effect size (OR = 5.63 (3.26-9.73)). These are cumulatively very rare, with a total allele count of only 169 spread across 71 different variants. As previously stated, these results exclude those for the nonsynonymous variant rs63750082, for which the alternate allele was observed in 62 cases and 5 controls.

With *SORL1* there is only weak evidence for association using the unweighted scores. Both LOF and splice region variants are rare and are associated with modest effect sizes.

Finally, the situation with *TREM2* is that the LOF variants are too rare to provide any evidence for association but that variants scoring highly with the MetaLR predictor produce weak evidence for association with SLP = 1.30 and a moderate effect with OR = 2.40 (0.98-5.84). They are rare, with a cumulative allele count of 393 spread over 29 separate variants. These results exclude rs75932628, for which the alternate allele was observed in 157 cases and 152 controls (OR = 1.71 (1.35 - 2.18)), and rs143332484, for which the alternate allele was observed in 290 cases and 419 controls (OR = 1.28 (1.09 - 1.50)).

Table 3 presents results for genes which did not produces statistically significant evidence for association in the current study but which were previously reported to be associated with AD and/or dementia. It can be seen that of these ten genes only two, *ADAM10* and *GRN*, show any evidence for association in the WGS dataset, with MLPs of 3.09 and 3.03 respectively. The results of analysing raw counts of different categories of variant in these genes are shown in Table 4. For *ADAM10*, this shows modest evidence for association of variants with high AM_prediction scores (SLP = 2.11) and the raw frequencies are more than twice as high among cases although the regression analysis including covariates estimates only a low effect size with OR = 1.11 (1.03-1.20). For all other genes, there is no real suggestion that any variant category provides evidence for association with AD.

**Table 3.**
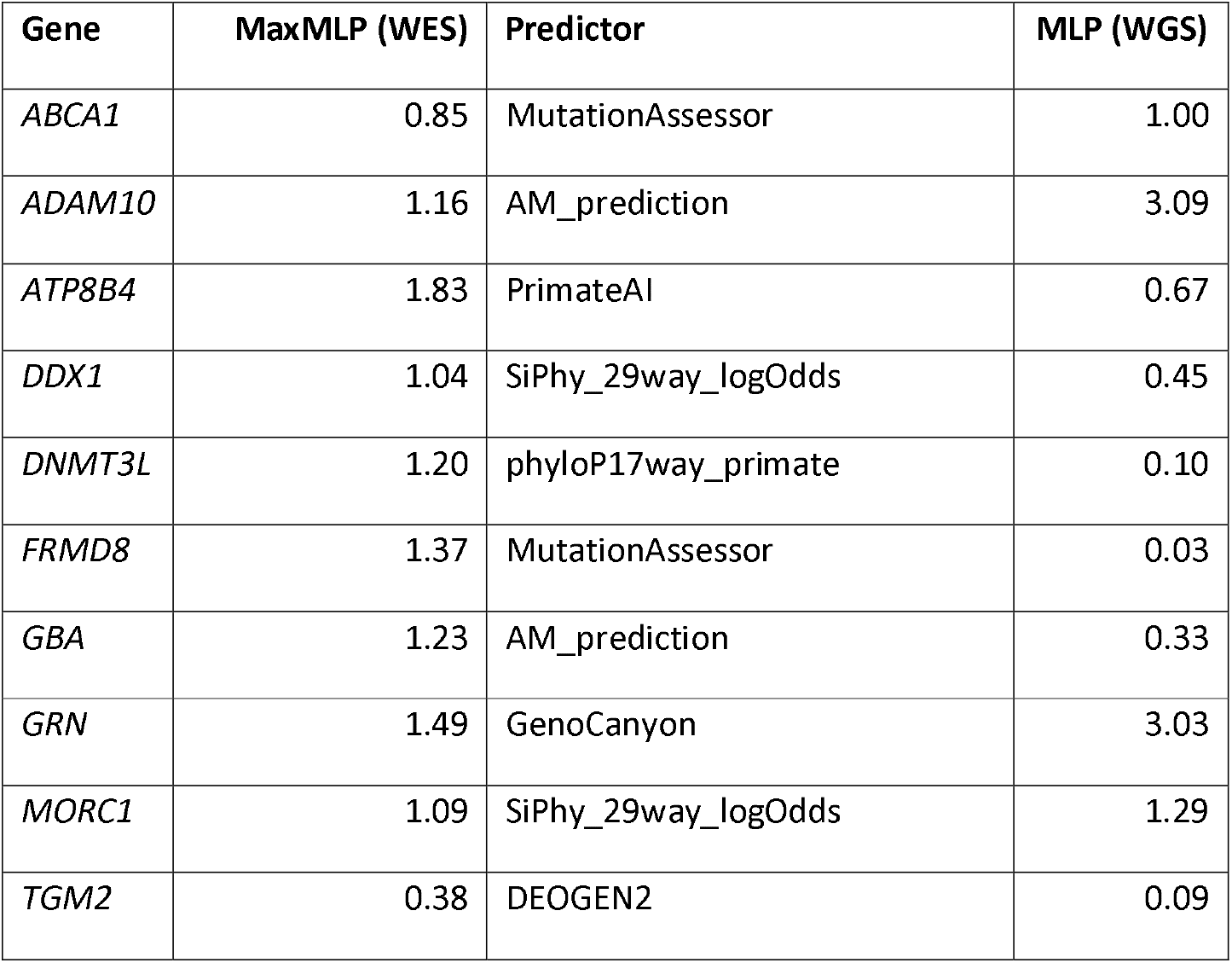
Results for other genes previously reported to be associated with AD and/or dementia. The table shows the maximum −log10(p) (MLP) produced by any of 45 pathogenicity predictors in the first stage analysis of exome-sequenced participants along with the predictor producing this MLP and the results obtained in the second stage using 40,983 participants. All results are obtained from multiple logistic regression analyses of AD with sex, 20 principal components and *APOE* genotypes as covariates. For all genes the MAF-weighted LOF scores and pathogenicity scores were both included in the analysis.

| Gene | MaxMLP (WES) | Predictor | MLP (WGS) |
| --- | --- | --- | --- |
| *ABCA1* | 0.85 | MutationAssessor | 1.00 |
| *ADAM10* | 1.16 | AM_prediction | 3.09 |
| *ATP8B4* | 1.83 | PrimateAI | 0.67 |
| *DDX1* | 1.04 | SiPhy_29way_logOdds | 0.45 |
| *DNMT3L* | 1.20 | phyloP17way_primate | 0.10 |
| *FRMD8* | 1.37 | MutationAssessor | 0.03 |
| *GBA* | 1.23 | AM_prediction | 0.33 |
| *GRN* | 1.49 | GenoCanyon | 3.03 |
| *MORC1* | 1.09 | SiPhy_29way_logOdds | 1.29 |
| *TGM2* | 0.38 | DEOGEN2 | 0.09 |

**Table 4.** Table showing results of multiple logistic regression analysis of AD in 40,983 participants for previously reported genes using raw counts of LOF and protein altering variants as well as those variants having a score of > 0.35 for the relevant predictor.

| Gene / category / predictor | Number of separate variants | Total count in controls | Mean count in controls | Total count in cases | Mean count in cases | OR (95% confidence interval) | SLP |
| --- | --- | --- | --- | --- | --- | --- | --- |
| *ABCA1* |  |  |  |  |  |  |  |
| LOF | 48 | 280 | 0.010104 | 126 | 0.009533 | 1.01 (0.79-1.28) | 0.02 |
| Protein altering | 493 | 3257 | 0.117473 | 1397 | 0.105638 | 1.06 (0.93-1.21) | 0.46 |
| MutationAssessor | 278 | 974 | 0.035141 | 418 | 0.031616 | 0.98 (0.73-1.33) | -0.04 |
| *ADAM10* |  |  |  |  |  |  |  |
| LOF | 10 | 7 | 0.000253 | 6 | 0.000454 | 2.90 (0.82-10.31) | 1.03 |
| Protein altering | 103 | 276 | 0.009950 | 172 | 0.013005 | 1.22 (0.98-1.52) | 1.19 |
| AM_prediction | 27 | 20 | 0.000721 | 22 | 0.001663 | 1.11 (1.03-1.20) | 2.11 |
| *ATP884* |  |  |  |  |  |  |  |
| LOF | 63 | 434 | 0.015643 | 219 | 0.016554 | 1.11 (0.91-1.35) | 0.55 |
| Protein altering | 318 | 3381 | 0.121862 | 1763 | 0.133250 | 1.09 (0.98-1.21) | 0.95 |
| PrimateAI | 58 | 1596 | 0.057526 | 863 | 0.065228 | 0.94 (0.75-1.18) | -0.23 |
| *DDX1* |  |  |  |  |  |  |  |
| LOF | 4 | 10 | 0.000360 | 3 | 0.000227 | 0.63 (0.16-2.51) | -0.30 |
| Protein altering | 126 | 275 | 0.009911 | 130 | 0.009825 | 1.26 (0.97-1.64) | 1.12 |
| SiPhy_29way_logOdds | 117 | 258 | 0.009299 | 105 | 0.007936 | 0.91 (0.81-1.01) | -1.12 |
| *DNMT3L* |  |  |  |  |  |  |  |
| LOF | 15 | 20 | 0.000721 | 11 | 0.000832 | 1.04 (0.48-2.26) | 0.03 |
| Protein altering | 88 | 355 | 0.012796 | 161 | 0.012170 | 0.97 (0.74-1.27) | -0.09 |
| phyloP17way_primate | 24 | 106 | 0.003821 | 63 | 0.004762 | 0.86 (0.36-2.03) | -0.14 |
| *FRMD8* |  |  |  |  |  |  |  |
| LOF | 16 | 15 | 0.000541 | 7 | 0.000529 | 0.88 (0.37-2.08) | -0.12 |
| Protein altering | 170 | 802 | 0.028923 | 393 | 0.029728 | 0.97 (0.84-1.12) | -0.19 |
| MutationAssessor | 26 | 142 | 0.005120 | 64 | 0.004838 | 1.15 (0.70-1.87) | 0.24 |
| *GBA* |  |  |  |  |  |  |  |
| LOF | 16 | 21 | 0.000759 | 6 | 0.000455 | 0.65 (0.25-1.70) | -0.43 |
| Protein altering | 137 | 1359 | 0.049100 | 742 | 0.056267 | 1.06 (0.96-1.17) | 0.57 |
| AM_prediction | 41 | 177 | 0.006381 | 80 | 0.006048 | 1.01 (0.97-1.05) | 0.12 |
| *GRN* |  |  |  |  |  |  |  |
| LOF | 32 | 619 | 0.022467 | 318 | 0.024172 | 1.13 (0.90-1.42) | 0.57 |
| Protein altering | 164 | 996 | 0.035932 | 464 | 0.035101 | 1.05 (0.87-1.27) | 0.24 |
| GenoCanyon | 106 | 956 | 0.034489 | 450 | 0.034040 | 1.05 (0.76-1.45) | 0.11 |
| *MORC1* |  |  |  |  |  |  |  |
| LOF | 32 | 61 | 0.002199 | 15 | 0.001134 | 0.51 (0.28-0.92) | -1.64 |
| Protein altering | 178 | 1124 | 0.040511 | 499 | 0.037714 | 0.95 (0.83-1.08) | -0.40 |
| SiPhy_29way_logOdds | 78 | 587 | 0.021156 | 269 | 0.020331 | 1.05 (0.92-1.20) | 0.33 |
| *TGM2* |  |  |  |  |  |  |  |
| LOF | 24 | 85 | 0.003065 | 48 | 0.003629 | 1.13 (0.77-1.67) | 0.29 |
| Protein altering | 232 | 1407 | 0.050715 | 667 | 0.050419 | 1.15 (0.93-1.42) | 0.70 |
| DEOGEN2 | 68 | 1170 | 0.042172 | 543 | 0.041044 | 0.85 (0.59-1.23) | -0.41 |

## Discussion

This association study of AD with rare coding variants weighted by rarity and predicted pathogenicity implicates four genes at conventional levels of statistical significance after correction for multiple testing: *ABCA7, PSEN1, SORL1* and *TREM2*. All of these genes have previously been reported to affect risk of AD, though it is worth noting that we show that extremely rare variants in *TREM2* with high MetaLR scores collectively contribute to risk in addition to the somewhat less rare variants which have previously been identified, rs75932628 and rs143332484. Of ten other previously reported AD genes, only *ADAM10* and *GRN2* produce any evidence for association using the methods applied here and analyses based on raw counts of variants do not provide any additional support. Consideration should be given as to whether the other previously reported genes might represent false positive findings.

Detailed consideration of the effects related to different categories of variant shows that there are 71 variants in *PSEN1* which have high scores with the phyloP17way_primate predictor (which is based on conservation scores) [23] and that these on average have a large effect on risk with OR = 5.63. However they are collectively rare with a mean count in cases of 0.00763, implying that only around 0.8% of cases might carry one of these variants. (This excludes consideration of the previously reported variant rs63750082, which was carried by 62 cases.) Although the other genes do produce statistically significant evidence for association, the estimated effect sizes are considerably lower. For *TREM2*, variants with a high MetalLR score have OR 2.40 but for *ABCA7* and *SORL1* no variant category has OR greater than 1.5 [24].

A potential benefit of understanding rare variant contributions to AD would be that they could be used to refine risk estimates for individuals and that this would have utility if and when interventions became available to prevent the development of AD. However the rarity and small effect sizes of risk variants in *ABCA7* and *SORL1* seem to make it unlikely that detecting them would be helpful in clinical practice. With regard to *TREM2*, one might consider genotyping rs75932628, which could potentially impact a decision to start treatment, but the risk allele would be seen in fewer than 0.5% of people. It does not seem likely that it would be worthwhile to use sequencing studies to detect other much rarer variants. By comparison, the *APOE* e4 allele has a greater effect on risk and is far commoner. Arguably, detection of a variant in *PSEN1* associated with a very large increase in risk might be worthwhile. However such variants would be seen in fewer than 1% of AD cases. Whether efforts should be made to detect them would depend on the availability of an effective preventative intervention and the costs and benefits associated with that intervention. For example, if an effective intervention were quite costly then one might only apply it to an individual at very high risk and then possibly identifying carriers of a *PSEN1* risk variant would enable targeted treatment. Overall though, the findings of this study do not suggest that there will be a large role for the identification of rare coding variants in the quantification of AD risk for predictive purposes. Their contribution to risk is dwarfed by that of *APOE*. Rather, the main benefit of these findings may be to throw further light on the pathogenesis of AD, with the ultimate aim being to support the development of novel therapeutic approaches.

## Data availability statement

The data used for this study is available on application to NIAGADS: https://www.niagads.org/.

## Code availability statement

Scripts, supporting files, interim files and full results will be deposited at the NIAGADS site: https://www.niagads.org/ and scripts are available at https://github.com/davenomiddlenamecurtis/ADSP2025scripts.

## Ethical approval

Ethical approval and informed consent had been obtained by the researchers who generated this dataset and ethics approval for the analyses performed was obtained from the UCL Research Ethics Committee (11527/003).

## Author contribution statement

DC obtained the data, carried out the analyses and wrote up the report. SJ contributed to the literature and highlighted important prior findings to inform the analytic approaches to be applied. The ADNI generated the dataset and made it available for researchers to access.

## Acknowledgments

The author wishes to acknowledge the staff supporting the High Performance Computing Cluster, Computer Science Department, University College London.

Data collection and sharing for the Alzheimer’s Disease Sequencing Project was funded by the Alzheimer’s Disease Neuroimaging Initiative (ADNI) (National Institutes of Health Grant U01 AG024904) and DOD ADNI (Department of Defense award number W81XWH-12-2-0012). ADNI is funded by the National Institute on Aging, the National Institute of Biomedical Imaging and Bioengineering, and through generous contributions from the following: AbbVie, Alzheimer’s Association; Alzheimer’s Drug Discovery Foundation; Araclon Biotech; BioClinica, Inc.; Biogen; Bristol-Myers Squibb Company; CereSpir, Inc.; Cogstate; Eisai Inc.; Elan Pharmaceuticals, Inc.; Eli Lilly and Company; EuroImmun; F. Hoffmann-La Roche Ltd and its affiliated company Genentech, Inc.; Fujirebio; GE Healthcare; IXICO Ltd.; Janssen Alzheimer Immunotherapy Research & Development, LLC.; Johnson & Johnson Pharmaceutical Research & Development LLC.; Lumosity; Lundbeck; Merck & Co., Inc.; Meso Scale Diagnostics, LLC.; NeuroRx Research; Neurotrack Technologies; Novartis Pharmaceuticals Corporation; Pfizer Inc.; Piramal Imaging; Servier; Takeda Pharmaceutical Company; and Transition Therapeutics. The Canadian Institutes of Health Research is providing funds to support ADNI clinical sites in Canada. Private sector contributions are facilitated by the Foundation for the National Institutes of Health (www.fnih.org). The grantee organization is the Northern California Institute for Research and Education, and the study is coordinated by the Alzheimer’s Therapeutic Research Institute at the University of Southern California. ADNI data are disseminated by the Laboratory for Neuro Imaging at the University of Southern California.

Data for this study were prepared, archived, and distributed by the National Institute on Aging Alzheimer’s Disease Data Storage Site (NIAGADS) at the University of Pennsylvania (U24-AG041689), funded by the National Institute on Aging. sa000001 - Alzheimer’s Disease Sequencing Project

The Alzheimer’s Disease Sequencing Project (ADSP) is comprised of two Alzheimer’s Disease (AD) genetics consortia and three National Human Genome Research Institute (NHGRI) funded Large Scale Sequencing and Analysis Centers (LSAC). The two AD genetics consortia are the Alzheimer’s Disease Genetics Consortium (ADGC) funded by NIA (U01 AG032984), and the Cohorts for Heart and Aging Research in Genomic Epidemiology (CHARGE) funded by NIA (R01 AG033193), the National Heart, Lung, and Blood Institute (NHLBI), other National Institute of Health (NIH) institutes and other foreign governmental and non-governmental organizations. The Discovery Phase analysis of sequence data is supported through UF1AG047133 (to Drs. Schellenberg, Farrer, Pericak-Vance, Mayeux, and Haines); U01AG049505 to Dr. Seshadri; U01AG049506 to Dr. Boerwinkle; U01AG049507 to Dr. Wijsman; and U01AG049508 to Dr. Goate and the Discovery Extension Phase analysis is supported through U01AG052411 to Dr. Goate, U01AG052410 to Dr. Pericak-Vance and U01 AG052409 to Drs. Seshadri and Fornage.

Sequencing for the Follow Up Study (FUS) is supported through U01AG057659 (to Drs. PericakVance, Mayeux, and Vardarajan) and U01AG062943 (to Drs. Pericak-Vance and Mayeux). Data generation and harmonization in the Follow-up Phase is supported by U54AG052427 (to Drs. Schellenberg and Wang). The FUS Phase analysis of sequence data is supported through U01AG058589 (to Drs. Destefano, Boerwinkle, De Jager, Fornage, Seshadri, and Wijsman), U01AG058654 (to Drs. Haines, Bush, Farrer, Martin, and Pericak-Vance), U01AG058635 (to Dr. Goate), RF1AG058066 (to Drs. Haines, Pericak-Vance, and Scott), RF1AG057519 (to Drs. Farrer and Jun), R01AG048927 (to Dr. Farrer), and RF1AG054074 (to Drs. Pericak-Vance and Beecham).

The ADGC cohorts include: Adult Changes in Thought (ACT) (U01 AG006781, U19 AG066567), the Alzheimer’s Disease Research Centers (ADRC) (P30 AG062429, P30 AG066468, P30 AG062421, P30 AG066509, P30 AG066514, P30 AG066530, P30 AG066507, P30 AG066444, P30 AG066518, P30 AG066512, P30 AG066462, P30 AG072979, P30 AG072972, P30 AG072976, P30 AG072975, P30 AG072978, P30 AG072977, P30 AG066519, P30 AG062677, P30 AG079280, P30 AG062422, P30 AG066511, P30 AG072946, P30 AG062715, P30 AG072973, P30 AG066506, P30 AG066508, P30 AG066515, P30 AG072947, P30 AG072931, P30 AG066546, P20 AG068024, P20 AG068053, P20 AG068077, P20 AG068082, P30 AG072958, P30 AG072959), the Chicago Health and Aging Project (CHAP) (R01 AG11101, RC4 AG039085, K23 AG030944), Indiana Memory and Aging Study (IMAS) (R01 AG019771), Indianapolis Ibadan (R01 AG009956, P30 AG010133), the Memory and Aging Project (MAP) (R01 AG17917), Mayo Clinic (MAYO) (R01 AG032990, U01 AG046139, R01 NS080820, RF1 AG051504, P50 AG016574), Mayo Parkinson’s Disease controls (NS039764, NS071674, 5RC2HG005605), University of Miami (R01 AG027944, R01 AG028786, R01 AG019085, IIRG09133827, A2011048), the Multi-Institutional Research in Alzheimer’s Genetic Epidemiology Study (MIRAGE) (R01 AG09029, R01 AG025259), the National Centralized Repository for Alzheimer’s Disease and Related Dementias (NCRAD) (U24 AG021886), the National Institute on Aging Late Onset Alzheimer’s Disease Family Study (NIA-LOAD) (U24 AG056270), the Religious Orders Study (ROS) (P30 AG10161, R01 AG15819), the Texas Alzheimer’s Research and Care Consortium (TARCC) (funded by the Darrell K Royal Texas Alzheimer’s Initiative), Vanderbilt University/Case Western Reserve University (VAN/CWRU) (R01 AG019757, R01 AG021547, R01 AG027944, R01 AG028786, P01 NS026630, and Alzheimer’s Association), the Washington Heights-Inwood Columbia Aging Project (WHICAP) (RF1 AG054023), the University of Washington Families (VA Research Merit Grant, NIA: P50AG005136, R01AG041797, NINDS: R01NS069719), the Columbia University Hispanic Estudio Familiar de Influencia Genetica de Alzheimer (EFIGA) (RF1 AG015473), the University of Toronto (UT) (funded by Wellcome Trust, Medical Research Council, Canadian Institutes of Health Research), and Genetic Differences (GD) (R01 AG007584). The CHARGE cohorts are supported in part by National Heart, Lung, and Blood Institute (NHLBI) infrastructure grant HL105756 (Psaty), RC2HL102419 (Boerwinkle) and the neurology working group is supported by the National Institute on Aging (NIA) R01 grant AG033193.

The CHARGE cohorts participating in the ADSP include the following: Austrian Stroke Prevention Study (ASPS), ASPS-Family study, and the Prospective Dementia Registry-Austria (ASPS/PRODEM-Aus), the Atherosclerosis Risk in Communities (ARIC) Study, the Cardiovascular Health Study (CHS), the Erasmus Rucphen Family Study (ERF), the Framingham Heart Study (FHS), and the Rotterdam Study (RS). ASPS is funded by the Austrian Science Fond (FWF) grant number P20545-P05 and P13180 and the Medical University of Graz. The ASPS-Fam is funded by the Austrian Science Fund (FWF) project I904), the EU Joint Programme – Neurodegenerative Disease Research (JPND) in frame of the BRIDGET project (Austria, Ministry of Science) and the Medical University of Graz and the Steiermärkische Krankenanstalten Gesellschaft. PRODEM-Austria is supported by the Austrian Research Promotion agency (FFG) (Project No. 827462) and by the Austrian National Bank (Anniversary Fund, project 15435. ARIC research is carried out as a collaborative study supported by NHLBI contracts (HHSN268201100005C, HHSN268201100006C, HHSN268201100007C, HHSN268201100008C, HHSN268201100009C, HHSN268201100010C, HHSN268201100011C, and HHSN268201100012C). Neurocognitive data in ARIC is collected by U01 2U01HL096812, 2U01HL096814, 2U01HL096899, 2U01HL096902, 2U01HL096917 from the NIH (NHLBI, NINDS, NIA and NIDCD), and with previous brain MRI examinations funded by R01-HL70825 from the NHLBI. CHS research was supported by contracts HHSN268201200036C, HHSN268200800007C, N01HC55222, N01HC85079, N01HC85080, N01HC85081, N01HC85082, N01HC85083, N01HC85086, and grants U01HL080295 and U01HL130114 from the NHLBI with additional contribution from the National Institute of Neurological Disorders and Stroke (NINDS). Additional support was provided by R01AG023629, R01AG15928, and R01AG20098 from the NIA. FHS research is supported by NHLBI contracts N01-HC-25195 and HHSN268201500001I. This study was also supported by additional grants from the NIA (R01s AG054076, AG049607 and AG033040 and NINDS (R01 NS017950). The ERF study as a part of EUROSPAN (European Special Populations Research Network) was supported by European Commission FP6 STRP grant number 018947 (LSHG-CT-2006-01947) and also received funding from the European Community’s Seventh Framework Programme (FP7/2007-2013)/grant agreement HEALTH-F4-2007-201413 by the European Commission under the programme “Quality of Life and Management of the Living Resources” of 5th Framework Programme (no. QLG2-CT-2002-01254). High-throughput analysis of the ERF data was supported by a joint grant from the Netherlands Organization for Scientific Research and the Russian Foundation for Basic Research (NWO-RFBR 047.017.043). The Rotterdam Study is funded by Erasmus Medical Center and Erasmus University, Rotterdam, the Netherlands Organization for Health Research and Development (ZonMw), the Research Institute for Diseases in the Elderly (RIDE), the Ministry of Education, Culture and Science, the Ministry for Health, Welfare and Sports, the European Commission (DG XII), and the municipality of Rotterdam. Genetic data sets are also supported by the Netherlands Organization of Scientific Research NWO Investments (175.010.2005.011, 911-03-012), the Genetic Laboratory of the Department of Internal Medicine, Erasmus MC, the Research Institute for Diseases in the Elderly (014-93-015; RIDE2), and the Netherlands Genomics Initiative (NGI)/Netherlands Organization for Scientific Research (NWO) Netherlands Consortium for Healthy Aging (NCHA), project 050-060-810. All studies are grateful to their participants, faculty and staff. The content of these manuscripts is solely the responsibility of the authors and does not necessarily represent the official views of the National Institutes of Health or the U.S. Department of Health and Human Services.

The FUS cohorts include: the Alzheimer’s Disease Research Centers (ADRC) (P30 AG062429, P30 AG066468, P30 AG062421, P30 AG066509, P30 AG066514, P30 AG066530, P30 AG066507, P30 AG066444, P30 AG066518, P30 AG066512, P30 AG066462, P30 AG072979, P30 AG072972, P30 AG072976, P30 AG072975, P30 AG072978, P30 AG072977, P30 AG066519, P30 AG062677, P30 AG079280, P30 AG062422, P30 AG066511, P30 AG072946, P30 AG062715, P30 AG072973, P30 AG066506, P30 AG066508, P30 AG066515, P30 AG072947, P30 AG072931, P30 AG066546, P20 AG068024, P20 AG068053, P20 AG068077, P20 AG068082, P30 AG072958, P30 AG072959), Alzheimer’s Disease Neuroimaging Initiative (ADNI) (U19AG024904), Amish Protective Variant Study (RF1AG058066), Cache County Study (R01AG11380, R01AG031272, R01AG21136, RF1AG054052), Case Western Reserve University Brain Bank (CWRUBB) (P50AG008012), Case Western Reserve University Rapid Decline (CWRURD) (RF1AG058267, NU38CK000480), CubanAmerican Alzheimer’s Disease Initiative (CuAADI) (3U01AG052410), Estudio Familiar de Influencia Genetica en Alzheimer (EFIGA) (5R37AG015473, RF1AG015473, R56AG051876), Genetic and Environmental Risk Factors for Alzheimer Disease Among African Americans Study (GenerAAtions) (2R01AG09029, R01AG025259, 2R01AG048927), Gwangju Alzheimer and Related Dementias Study (GARD) (U01AG062602), Hillblom Aging Network (2014-A-004-NET, R01AG032289, R01AG048234), Hussman Institute for Human Genomics Brain Bank (HIHGBB) (R01AG027944, Alzheimer’s Association “Identification of Rare Variants in Alzheimer Disease”), Ibadan Study of Aging (IBADAN) (5R01AG009956), Longevity Genes Project (LGP) and LonGenity (R01AG042188, R01AG044829, R01AG046949, R01AG057909, R01AG061155, P30AG038072), Mexican Health and Aging Study (MHAS) (R01AG018016), Multi-Institutional Research in Alzheimer’s Genetic Epidemiology (MIRAGE) (2R01AG09029, R01AG025259, 2R01AG048927), Northern Manhattan Study (NOMAS) (R01NS29993), Peru Alzheimer’s Disease Initiative (PeADI) (RF1AG054074), Puerto Rican 1066 (PR1066) (Wellcome Trust (GR066133/GR080002), European Research Council (340755)), Puerto Rican Alzheimer Disease Initiative (PRADI) (RF1AG054074), Reasons for Geographic and Racial Differences in Stroke (REGARDS) (U01NS041588), Research in African American Alzheimer Disease Initiative (REAAADI) (U01AG052410), the Religious Orders Study (ROS) (P30 AG10161, P30 AG72975, R01 AG15819, R01 AG42210), the RUSH Memory and Aging Project (MAP) (R01 AG017917, R01 AG42210Stanford Extreme Phenotypes in AD (R01AG060747), University of Miami Brain Endowment Bank (MBB), University of Miami/Case Western/North Carolina A&T African American (UM/CASE/NCAT) (U01AG052410, R01AG028786), Wisconsin Registry for Alzheimer’s Prevention (WRAP) (R01AG027161 and R01AG054047), Mexico-Southern California Autosomal Dominant Alzheimer’s Disease Consortium (R01AG069013), Center for Cognitive Neuroscience and Aging (R01AG047649), and the A4 Study (R01AG063689, U19AG010483 and U24AG057437).

The four LSACs are: the Human Genome Sequencing Center at the Baylor College of Medicine (U54 HG003273), the Broad Institute Genome Center (U54HG003067), The American Genome Center at the Uniformed Services University of the Health Sciences (U01AG057659), and the Washington University Genome Institute (U54HG003079). Genotyping and sequencing for the ADSP FUS is also conducted at John P. Hussman Institute for Human Genomics (HIHG) Center for Genome Technology (CGT).

Biological samples and associated phenotypic data used in primary data analyses were stored at Study Investigators institutions, and at the National Centralized Repository for Alzheimer’s Disease and Related Dementias (NCRAD, U24AG021886) at Indiana University funded by NIA. Associated Phenotypic Data used in primary and secondary data analyses were provided by Study Investigators, the NIA funded Alzheimer’s Disease Centers (ADCs), and the National Alzheimer’s Coordinating Center (NACC, U24AG072122) and the National Institute on Aging Genetics of Alzheimer’s Disease Data Storage Site (NIAGADS, U24AG041689) at the University of Pennsylvania, funded by NIA. Harmonized phenotypes were provided by the ADSP Phenotype Harmonization Consortium (ADSP-PHC), funded by NIA (U24 AG074855, U01 AG068057 and R01 AG059716) and Ultrascale Machine Learning to Empower Discovery in Alzheimer’s Disease Biobanks (AI4AD, U01 AG068057). This research was supported in part by the Intramural Research Program of the National Institutes of health, National Library of Medicine. Contributors to the Genetic Analysis Data included Study Investigators on projects that were individually funded by NIA, and other NIH institutes, and by private U.S. organizations, or foreign governmental or nongovernmental organizations.

The ADSP Phenotype Harmonization Consortium (ADSP-PHC) is funded by NIA (U24 AG074855, U01 AG068057 and R01 AG059716). The harmonized cohorts within the ADSP-PHC include:lllthe Anti-Amyloid Treatment in Asymptomatic Alzheimer’s study (A4 Study), a secondary prevention trial in preclinical Alzheimer’s disease, aiming to slow cognitive decline associated with brain amyloid accumulation in clinically normal older individuals. The A4 Study is funded by a public-private-philanthropic partnership, including funding from the National Institutes of Health-National Institute on Aging, Eli Lilly and Company, Alzheimer’s Association, Accelerating Medicines Partnership, GHR Foundation, an anonymous foundation and additional private donors, with in-kind support from Avid and Cogstate. The companion observational Longitudinal Evaluation of Amyloid Risk and Neurodegeneration (LEARN) Study is funded by the Alzheimer’s Association and GHR Foundation. The A4 and LEARN Studies are led by Dr. Reisa Sperling at Brigham and Women’s Hospital, Harvard Medical School and Dr. Paul Aisen at the Alzheimer’s Therapeutic Research Institute (ATRI), University of Southern California. The A4 and LEARN Studies are coordinated by ATRI at the University of Southern California, and the data are made available through the Laboratory for Neuro Imaging at the University of Southern California. The participants screening for the A4 Study provided permission to share their de-identified data in order to advance the quest to find a successful treatment for Alzheimer’s disease. We would like to acknowledge the dedication of all the participants, the site personnel, and all of the partnership team members who continue to make the A4 and LEARN Studies possible. The complete A4 Study Team list is available on: a4study.org/a4-study-team.; the Adult Changes in Thought study (ACT), Alzheimer’s Disease Neuroimaging Initiative (ADNI): *Data collection and sharing for this project was funded by the Alzheimer’s Disease Neuroimaging Initiative (ADNI) (National Institutes of Health Grant U01 AG024904) and DOD ADNI (Department of Defense award number W81XWH-12-2-0012). ADNI is funded by the National Institute on Aging, the National Institute of Biomedical Imaging and Bioengineering, and through generous contributions from the following: AbbVie, Alzheimer’s Association; Alzheimer’s Drug Discovery Foundation; Araclon Biotech; BioClinica, Inc*.; *Biogen; Bristol-Myers Squibb Company; CereSpir, Inc*.; *Cogstate; Eisai Inc*.; *Elan Pharmaceuticals, Inc*.; *Eli Lilly and Company; EuroImmun; F. Hoffmann-La Roche Ltd and its affiliated company Genentech, Inc*.; *Fujirebio; GE Healthcare; IXICO Ltd*.;*Janssen Alzheimer Immunotherapy Research & Development, LLC*.; *Johnson & Johnson Pharmaceutical Research & Development LLC*.; *Lumosity; Lundbeck; Merck & Co*., *Inc*.;*Meso Scale Diagnostics, LLC*.; *NeuroRx Research; Neurotrack Technologies; Novartis Pharmaceuticals Corporation; Pfizer Inc*.; *Piramal Imaging; Servier; Takeda Pharmaceutical Company; and Transition Therapeutics. The Canadian Institutes of Health Research is providing funds to support ADNI clinical sites in Canada. Private sector contributions are facilitated by the Foundation for the National Institutes of Health (www.fnih.org). The grantee organization is the Northern California Institute for Research and Education, and the study is coordinated by the Alzheimer’s Therapeutic Research Institute at the University of Southern California. ADNI data are disseminated by the Laboratory for Neuro Imaging at the University of Southern California; E*studio Familiar de Influencia Genetica en Alzheimer (EFIGA): *5R37AG015473, RF1AG015473, R56AG051876;* the Health & Aging Brain Study – Health Disparities (HABS-HD), supported by the National Institute on Aging of the National Institutes of Health under Award Numbers R01AG054073, R01AG058533, R01AG070862, P41EB015922, and U19AG078109; the Korean Brain Aging Study for the Early Diagnosis and Prediction of Alzheimer’s disease (KBASE), which was supported by a grant from Ministry of Science, ICT and Future Planning (Grant No: NRF-2014M3C7A1046042); Memory & Aging Project at Knight Alzheimer’s Disease Research Center (MAP at Knight ADRC): The Memory and Aging Project at the Knight-ADRC (Knight-ADRC). This work was supported by the National Institutes of Health (NIH) grants R01AG064614, R01AG044546, RF1AG053303, RF1AG058501, U01AG058922 and R01AG064877 to Carlos Cruchaga. The recruitment and clinical characterization of research participants at Washington University was supported by NIH grants P30AG066444, P01AG03991, and P01AG026276. Data collection and sharing for this project was supported by NIH grants RF1AG054080, P30AG066462, R01AG064614 and U01AG052410. We thank the contributors who collected samples used in this study, as well as patients and their families, whose help and participation made this work possible. This work was supported by access to equipment made possible by the Hope Center for Neurological Disorders, the Neurogenomics and Informatics Center (NGI: https://neurogenomics.wustl.edu/) and the Departments of Neurology and Psychiatry at Washington University School of Medicine; National Alzheimer’s Coordinating Center (NACC): *The NACC database is funded by NIA/NIH Grant U24 AG072122*. SCAN is a multi-institutional project that was funded as a U24 grant (AG067418) by the National Institute on Aging in May 2020. Data collected by SCAN and shared by NACC are contributed by the NIA-funded ADRCs as follows: *P30 AG062429 (PI James Brewer, MD, PhD), P30 AG066468 (PI Oscar Lopez, MD), P30 AG062421 (PI Bradley Hyman, MD, PhD), P30 AG066509 (PI Thomas Grabowski, MD), P30 AG066514 (PI Mary Sano, PhD), P30 AG066530 (PI Helena Chui, MD), P30 AG066507 (PI Marilyn Albert, PhD), P30 AG066444 (PI John Morris, MD), P30 AG066518 (PI Jeffrey Kaye, MD), P30 AG066512 (PI Thomas Wisniewski, MD), P30 AG066462 (PI Scott Small, MD), P30 AG072979 (PI David Wolk, MD), P30 AG072972 (PI Charles DeCarli, MD), P30 AG072976 (PI Andrew Saykin, PsyD), P30 AG072975 (PI David Bennett, MD), P30 AG072978 (PI Neil Kowall, MD), P30 AG072977 (PI Robert Vassar, PhD), P30 AG066519 (PI Frank LaFerla, PhD), P30 AG062677 (PI Ronald Petersen, MD, PhD), P30 AG079280 (PI Eric Reiman, MD), P30 AG062422 (PI Gil Rabinovici, MD), P30 AG066511 (PI Allan Levey, MD, PhD), P30 AG072946 (PI Linda Van Eldik, PhD), P30 AG062715 (PI Sanjay Asthana, MD, FRCP), P30 AG072973 (PI Russell Swerdlow, MD), P30 AG066506 (PI Todd Golde, MD, PhD), P30 AG066508 (PI Stephen Strittmatter, MD, PhD), P30 AG066515 (PI Victor Henderson, MD, MS), P30 AG072947 (PI Suzanne Craft, PhD), P30 AG072931 (PI Henry Paulson, MD, PhD), P30 AG066546 (PI Sudha Seshadri, MD), P20 AG068024 (PI Erik Roberson, MD, PhD), P20 AG068053 (PI Justin Miller, PhD), P20 AG068077 (PI Gary Rosenberg, MD), P20 AG068082 (PI Angela Jefferson, PhD), P30 AG072958 (PI Heather Whitson, MD), P30 AG072959 (PI James Leverenz, MD)*; National Institute on Aging Alzheimer’s Disease Family Based Study (NIA-AD FBS): U24 AG056270; Religious Orders Study (ROS): P30AG10161,R01AG15819, R01AG42210; Memory and Aging Project (MAP - Rush): R01AG017917, R01AG42210; Minority Aging Research Study (MARS): R01AG22018, R01AG42210; the Texas Alzheimer’s Research and Care Consortium (TARCC), funded by the Darrell K Royal Texas Alzheimer’s Initiative, directed by the Texas Council on Alzheimer’s Disease and Related Disorders; Washington Heights/Inwood Columbia Aging Project (WHICAP): *RF1 AG054023;* and Wisconsin Registry for Alzheimer’s Prevention (WRAP): *R01AG027161 and R01AG054047*. Additional acknowledgments include the National Institute on Aging Genetics of Alzheimer’s Disease Data Storage Site (NIAGADS, U24AG041689) at the University of Pennsylvania, funded by NIA.

## References

1. Caplan LR, Simon RP, Hassani S. Cerebrovascular Disease. Neurobiology of Brain Disorders: Biological Basis of Neurological and Psychiatric Disorders, Second Edition [Internet]. 2023 Aug 7 [cited 2024 Aug 7];457–76. Available from: https://www.ncbi.nlm.nih.gov/books/NBK430927/

2. Latimer CS, Lucot KL, Keene CD, Cholerton B, Montine TJ. Genetic Insights into Alzheimer’s Disease. Annu Rev Pathol [Internet]. 2021 Jan 24 [cited 2024 Nov 8];16:351–76. Available from: https://pubmed.ncbi.nlm.nih.gov/33497263/

3. Kjeldsen EW, Frikke-Schmidt R. Causal cardiovascular risk factors for dementia-insights from observational and genetic studies. Cardiovasc Res [Internet]. 2024 Nov 5 [cited 2024 Nov 11]; Available from: https://pubmed.ncbi.nlm.nih.gov/39498825/

4. Curtis D, Bakaya K, Sharma L, Bandyopadhay S. Weighted burden analysis of exomesequenced late onset Alzheimer’s cases and controls provides further evidence for involvement of PSEN1 and demonstrates protective role for variants in tyrosine phosphatase genes. Ann Hum Genet [Internet]. 2019 Apr 2 [cited 2019 Apr 24];84:291–302. Available from: https://www.biorxiv.org/content/10.1101/596007v1

5. Jonsson T, Stefansson H, Steinberg S, Jonsdottir I, Jonsson P V., Snaedal J, et al. Variant of TREM2 associated with the risk of Alzheimer’s disease. New England Journal of Medicine. 2013 Jan 10;368(2):107–16.

6. Holstege H, Hulsman M, Charbonnier C, Grenier-Boley B, Quenez O, Grozeva D, et al. Exome sequencing identifies rare damaging variants in ATP8B4 and ABCA1 as risk factors for Alzheimer’s disease. Nature Genetics 2022 54:12 [Internet]. 2022 Nov 21 [cited 2024 Nov 8];54(12):1786–94. Available from: https://www.nature.com/articles/s41588-022-01208-7

7. Zhang YR, Wu BS, Chen SD, Yang L, Deng YT, Guo Y, et al. Whole exome sequencing analyses identified novel genes for Alzheimer’s disease and related dementia. Alzheimers Dement [Internet]. 2024 Oct 1 [cited 2024 Nov 8];20(10). Available from: https://pubmed.ncbi.nlm.nih.gov/39129223/

8. Gibbons L, Curtis D. Analysis of 470,000 exome-sequenced UK Biobank participants identifies genes containing rare variants which confer dementia risk. Journal of Neurogenetics [in press]. 2025;

9. Kuzma A, Valladares O, Greenfest-Allen E, Nicaretta H, Kirsch M, Ren Y, et al. NIAGADS: A Comprehensive National Data Repository for Alzheimer’s Disease and Related Dementia Genetics and Genomics Research. medRxiv [Internet]. 2024 Dec 7 [cited 2025 Mar 12];2024.10.07.24315029. Available from: https://www.medrxiv.org/content/10.1101/2024.10.07.24315029v3

10. Hurko O, Black SE, Doody R, Doraiswamy PM, Gamst A, Kaye J, et al. The ADNI Publication Policy: Commensurate recognition of critical contributors who are not authors. Vol. 59, NeuroImage. Neuroimage; 2012. p. 4196–200.

11. Beecham GW, Bis JC, Martin ER, Choi SH, DeStefano AL, van Duijn CM, et al. The Alzheimer’s Disease Sequencing Project: Study design and sample selection. Neurol Genet. 2017 Oct 13;3(5):e194.

12. NIAGADS. NG00067 (Version 2). https://dss.niagads.org/datasets/ng00067/. 2020.

13. Bis JC, Jian X, Kunkle BW, Chen Y, Hamilton-Nelson KL, Bush WS, et al. Whole exome sequencing study identifies novel rare and common Alzheimer’s-Associated variants involved in immune response and transcriptional regulation. Mol Psychiatry. 2018 Aug 14;1.

14. NIAGADS. NG00067 (Version 16). https://dss.niagads.org/datasets/ng00067/. 2025.

15. Leung YY, Lee WP, Kuzma AB, Nicaretta H, Valladares O, Gangadharan P, et al. Alzheimer’s Disease Sequencing Project Release 4 Whole Genome Sequencing Dataset. medRxiv [Internet]. 2024 Dec 6 [cited 2025 Jan 24];2024.12.03.24317000. Available from: https://www.medrxiv.org/content/10.1101/2024.12.03.24317000v1

16. McLaren W, Gil L, Hunt SE, Riat HS, Ritchie GRS, Thormann A, et al. The Ensembl Variant Effect Predictor. Genome Biol [Internet]. 2016 Jun 6 [cited 2017 May 9];17(1):122. Available from: http://genomebiology.biomedcentral.com/articles/10.1186/s13059-016-0974-4

17. Cheng J, Novati G, Pan J, Bycroft C, Žemgulytė A, Applebaum T, et al. Accurate proteome-wide missense variant effect prediction with AlphaMissense. Science (1979) [Internet]. 2023 Sep 22 [cited 2023 Nov 17];381(6664). Available from: https://pubmed.ncbi.nlm.nih.gov/37733863/

18. Liu X, Li C, Mou C, Dong Y, Tu Y. dbNSFP v4: a comprehensive database of transcript-specific functional predictions and annotations for human nonsynonymous and splice-site SNVs. Genome Med. 2020 Dec 1;12(1).

19. Curtis D. A rapid method for combined analysis of common and rare variants at the level of a region, gene, or pathway. Adv Appl Bioinform Chem. 2012;5:1–9.

20. Curtis D. Exploration of weighting schemes based on allele frequency and annotation for weighted burden association analysis of complex phenotypes. Gene [Internet]. 2022 Jan 30 [cited 2023 Aug 23];809. Available from: https://pubmed.ncbi.nlm.nih.gov/34688815/

21. Curtis D. Assessment of ability of AlphaMissense to identify variants affecting susceptibility to common disease. European Journal of Human Genetics 2024 [Internet]. 2024 Aug 3 [cited 2024 Aug 22];1–9. Available from: https://www.nature.com/articles/s41431-024-01675-y

22. Lee JH, Kahn A, Cheng R, Reitz C, Vardarajan B, Lantigua R, et al. Disease-related mutations among Caribbean Hispanics with familial dementia. Mol Genet Genomic Med [Internet]. 2014 Sep 1 [cited 2025 Feb 14];2(5):430–7. Available from: https://pubmed.ncbi.nlm.nih.gov/25333068/

23. Pollard KS, Hubisz MJ, Rosenbloom KR, Siepel A. Detection of nonneutral substitution rates on mammalian phylogenies. Genome Res [Internet]. 2010 Jan 1 [cited 2025 Mar 7];20(1):110–21. Available from: https://genome.cshlp.org/content/20/1/110.full

24. Dong C, Wei P, Jian X, Gibbs R, Boerwinkle E, Wang K, et al. Comparison and integration of deleteriousness prediction methods for nonsynonymous SNVs in whole exome sequencing studies. Hum Mol Genet [Internet]. 2015 Apr 15 [cited 2025 Mar 7];24(8):2125–37. Available from: 10.1093/hmg/ddu733

